# Multimodal ctDNA profiling for cancer detection and monitoring in pan-cancer patients with advanced disease enrolled in the SHIVA02 trial

**DOI:** 10.64898/2026.07.24.26358660

**Authors:** Kenza Nedara, Marine Gorse, Klaus von Grafenstein, Julien Masliah-Planchon, Samantha Antonio, Charline Bianchi, Mathieu Séné, Pauline du Rusquec, Odette Mariani, Maud Kamal, Abderaouf Hamza, Ivan Bièche, Christophe Le Tourneau, Célia Dupain, Charlotte Proudhon

## Abstract

**Background:** Liquid biopsy-based monitoring of circulating tumor DNA (ctDNA) holds promises for real-time assessment of tumor burden and treatment response in precision oncology. However, mutation-based approaches alone show limited sensitivity, particularly in low-shedding tumors. We evaluated whether integrating tumor-naive epigenomic biomarkers — specifically LINE-1 retrotransposon (L1PA) hypomethylation and copy number variation (CNV) — with standard mutation-based ctDNA analysis could improve cancer detection and longitudinal monitoring in patients enrolled in the SHIVA02 precision oncology trial.

**Methods:** We performed a retrospective analysis of 32 patients with 11 different types of advanced or metastatic solid tumors and who received molecularly matched targeted therapies within the SHIVA02 trial (NCT03084757). Plasma samples were collected at baseline, longitudinally every two months and at progression. Multimodal ctDNA profiling was performed using the DRAGON targeted NGS panel covering SNVs, indels and focal CNVs in 571 genes and the DIAMOND assay profiling L1PA methylation and genome-wide CNV. A three-step classification algorithm integrating maximum variant allele frequency (MaxVAF), L1PA methylation-based cancer probability (MethP_Cancer_), and genome-wide CNV scores was developed to maximize ctDNA detectability.

**Results:** At baseline, mutation-based profiling detected ctDNA in 62.5% of patients. L1PA hypomethylation alone identified ctDNA in 90.6% of patients, including cases with undetectable mutations. The three-step integrative model increased overall detectability to 96.9%. The three modalities showed limited pairwise correlation at baseline, supporting their complementarity. Longitudinal analysis demonstrated that changes in MaxVAF, MethP_Cancer_, and L1PA CNV scores over time were informative on treatment response across tumor types, with ctDNA detected in 97.3% of samples collected at disease progression. In selected patients, ctDNA alterations preceded radiological progression by two to four months.

**Conclusions:** Multimodal ctDNA profiling integrating mutation analysis, CNV profiling, and tumor-naive LINE-1 hypomethylation substantially improves ctDNA detection at baseline and during treatment in a pan-cancer setting. These complementary genomic and epigenomic biomarkers provide a more comprehensive and dynamic assessment of tumor burden than single-modality approaches, supporting prospective validation in larger cohorts for integration into precision oncology workflows.

## Introduction

The advent of precision oncology has profoundly transformed cancer care by enabling therapeutic decisions guided not only by tumor histology but also by molecular alterations that drive oncogenesis [1], [2], [3]. The integration of high-throughput sequencing into clinical workflows have broadened access to individualized treatment strategies. In this context, molecular tumor boards have emerged as essential structures to interpret increasingly complex genomic data and optimize treatment recommendations according to clinical actionability frameworks [4]. Large-scale precision medicine trials, including MOSCATO-01, SHIVA01, WINTHER, and more recently ROME have demonstrated the feasibility of implementing comprehensive molecular profiling and molecular tumor board-guided treatment strategies in patients with advanced cancers. Collectively, these studies support the clinical relevance of molecularly guided therapies but reveal that only a small proportion of patients ultimately received targeted therapies and that clinical benefit among treated patients was heterogeneous [5], [6], [7], [8]. To address interpatient heterogeneity, alternative precision oncology trial designs have been developed in which patients serve as their own controls, using the progression free survival (PFS) ratio as a measure of individual benefit. In MOSCATO-01 and WINTHER the PFS ratio relied on a retrospectively determined PFS1, introducing potential bias when the timing and modalities of tumor assessments differed between treatment lines. The SHIVA02 trial (NCT03084757), a tissue-agnostic precision medicine study in advanced and/or metastatic cancers and the first to prospectively assess both PFS1 and PFS2, showed that matched therapy was achieved in 13% of patients, among whom 32% experienced a PFS ratio above 1.3 [9].

Traditionally, molecular profiling relies on surgical specimens or tissue biopsies. However, tissue-based approaches present several limitations, including procedural invasiveness, sampling bias, and an inability to fully capture intratumor heterogeneity or emerging resistance mechanisms during treatment. Moreover, repeated tissue sampling is often not feasible in patients with advanced or metastatic disease because of clinical deterioration and/or limited lesion accessibility. To overcome these constraints, we have previously shown that less invasive procedures, such as fine-needle aspiration, can reliably detect actionable genomic alterations, underscoring the need to explore alternative sampling strategies [10], [11]. Among these alternative strategies, liquid biopsy has emerged as a minimally invasive and dynamic tool capable of providing real-time molecular insights.

Circulating tumor DNA (ctDNA) has emerged as a particularly promising biomarker reflecting tumor burden, clonal evolution, and treatment-induced changes [12], [13]. Longitudinal monitoring of ctDNA has demonstrated the ability to detect disease progression earlier than radiological imaging across multiple tumor types as shown in large prospective studies and disease-specific clinical cohorts [14], [15], [16]. Representative examples include *EGFR*- mutated non–small cell lung cancer, where early ctDNA progression precedes radiologic confirmation [17]; metastatic breast cancer, where ctDNA tracing informs response and acquired resistance mechanisms [17], [18], [19], [20], [21]; and metastatic colorectal cancer, in which ctDNA progression anticipates radiological progression in more than half of patients [22]. Recent reviews reinforce the clinical utility of ctDNA in precision oncology, particularly for response assessment and personalized therapeutic strategies [23], [24]. However, mutation-based ctDNA analyses may be limited in low-shedding tumors, highlighting the need for complementary biomarkers to improve sensitivity [13]. Beyond genomic alterations, epigenetic modifications — including DNA methylation changes — constitute early, widespread, and stable hallmarks of cancer. These features are preserved in cell-free DNA and can provide sensitive tumor-derived signals, even in settings where ctDNA mutational burden is low. Methylation-based ctDNA assays have shown potential for early cancer detection, minimal residual disease assessment, treatment outcome prediction, and tissue-of-origin inference [25], [26], [27], [28]. Among these epigenetic biomarkers, LINE-1 reactivation represents a global indicator of cancer-associated epigenomic deregulation. We recently demonstrated that LINE-1 hypomethylation and copy number variations (CNV) detected from plasma DNA can reliably discriminate cancer patients with multiple cancer types from healthy individuals [29]. This approach could complement mutation-based ctDNA detection strategies, thereby enhancing the sensitivity of liquid biopsy approaches.

In this context, we conducted a retrospective analysis of 32 patients, enrolled in the SHIVA02 trial (NCT03084757), who received a targeted therapy or immunotherapy based on their molecular profile and followed with liquid biopsies. The objectives of our study were to: (1) evaluate the ability of ctDNA mutational profiling to detect molecular alterations across tumor types relative to core-needle biopsy; (2) assess correlations between LINE-1 hypomethylation, CNV and genomic alterations; (3) characterize the dynamic evolution of these biomarkers during treatment and determine their associations with therapeutic response; and (4) set up an algorithm to automatically detect ctDNA-containing samples.

## Results

### The SHIVA02 trial: molecular profiling-based targeted therapy including non-invasive disease monitoring using plasma DNA

The cohort in our study included 32 patients enrolled as part of the SHIVA02 precision-oncology trial who were molecularly oriented to matched therapy. Patients first received conventional treatment (Treatment 1, Step 1) while undergoing molecular profiling on baseline tumor biopsies (**Figure 1A**). When patients progressed on Treatment 1, they were then included in the second phase of the trial (Step 2) and received matched therapy tailored to the mutation identified from their tumor (Treatment 2). Baseline tumor samples were collected for all patients at inclusion, while plasma samples were obtained at baseline, every two months and at progression throughout Step 1 and Step 2 (**Figure 1A-B**, [9]).

**Figure 1.**
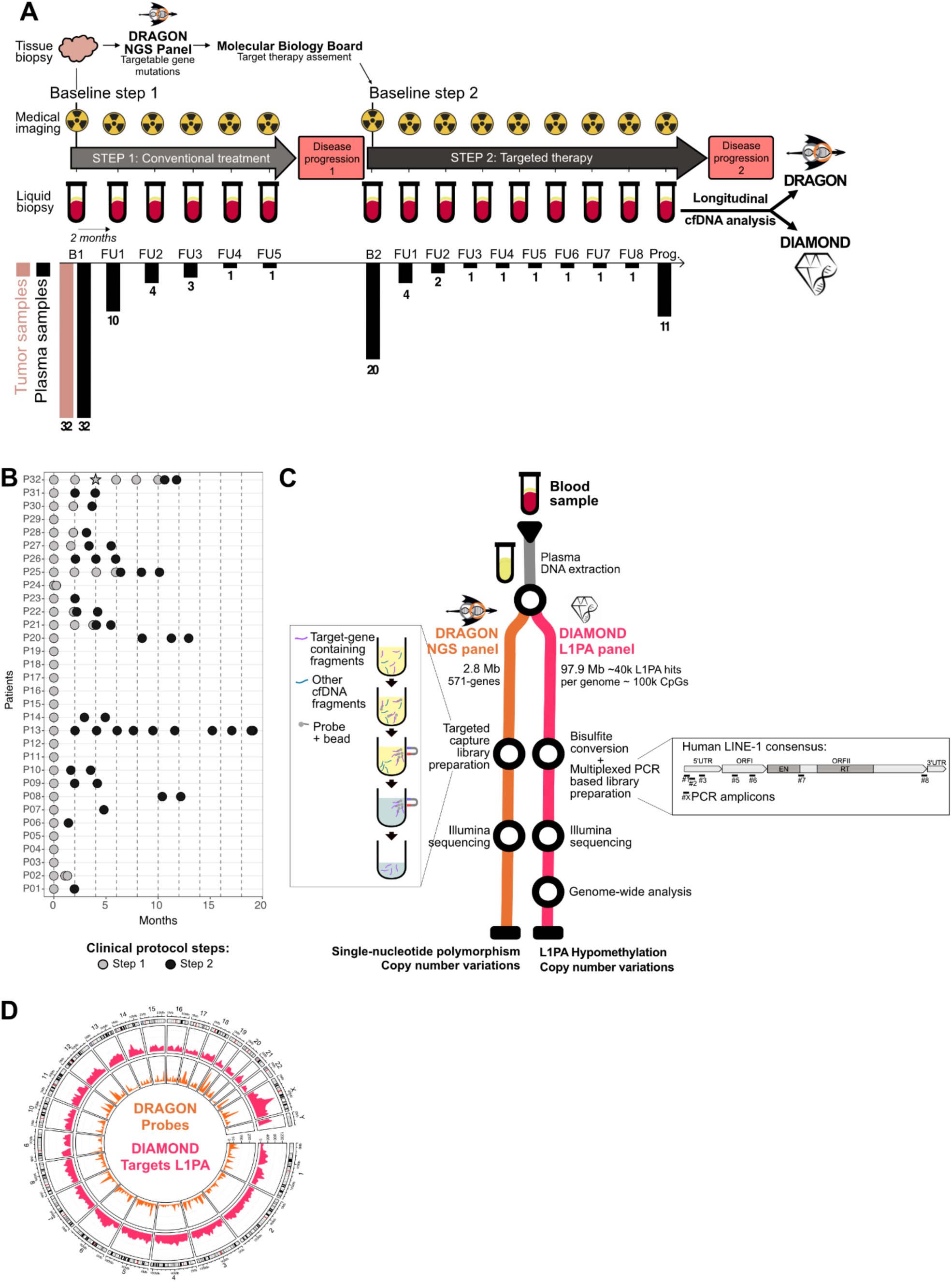
The SHIVA02 trial: molecular profiling-based targeted therapy including non-invasive disease monitoring using plasma DNA. **A.** Design of the SHIVA02 study for patients molecularly oriented to targeted therapy and description of paired tumor / plasma samples collected at each time point. Histograms and numbers indicate the number of patients with sampling at each follow up time point. Plasma DNA longitudinal analysis was performed using a custom 571-gene panel: DRAGON, and the L1PA-targeted bisulfite sequencing method: DIAMOND. FU: Follow up time point. **B.** Blood sampling points over time, treatment steps and molecular methods used for profiling each patient. Circles: DRAGON + DIAMOND profiling, Star: DRAGON only. **C.** Description of the DRAGON and DIAMOND methods used for molecular characterization. **D.** Circos plot showing the distribution of the DRAGON probes (orange) and L1PA hits targeted by DIAMOND (pink) on the hg38 genome.

The cohort spans 11 types of cancers (**Table S1**). The most frequent tumor types represented in the cohort were breast (31%), gastrointestinal (31%), and gynecological (28%) cancers. Consequently, the cohort was predominantly female (88%) with a median age of 60 years (**Table 1**). During Step 1, based on best overall response according to RECIST criteria, 75% of patients experienced disease progression, 19% achieved stable disease, and 6% showed a partial response as best response (**Table 1**). Most frequent targeted therapies received at Step 2 were BRAF/MEK inhibitors, PI3K inhibitors, and CDK4/6 inhibitors, as summarized in **Table 1**. Outcomes and response assessment after and during Step 2, using the same RECIST criteria, showed a similar distribution, with 72% progression within the first 2 months, 22% stable disease, and 3% partial response (**Table 1**,[9]).

**Table 1.**
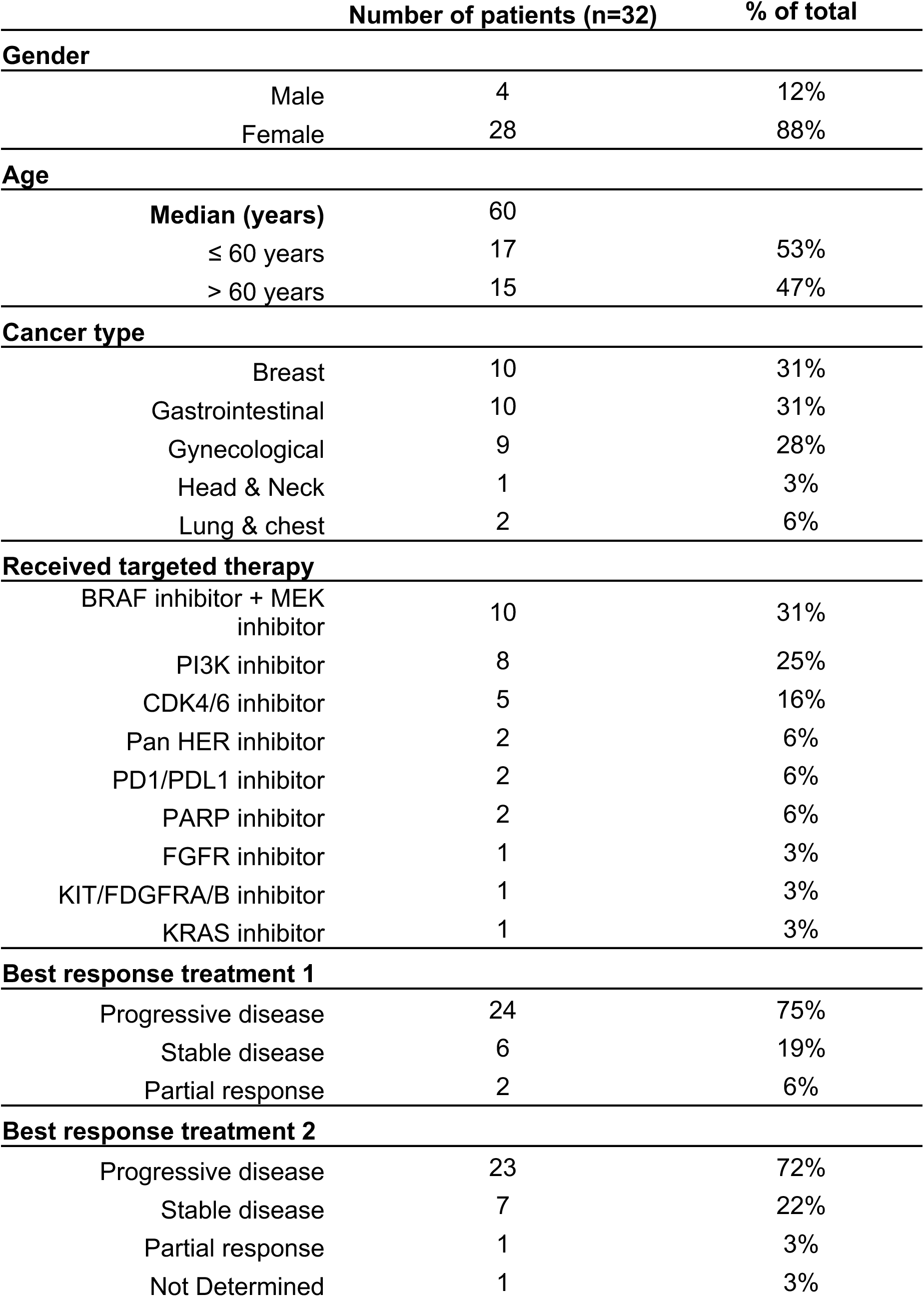
Characteristics of SHIVA02 patients receiving targeted therapy.

Molecular profiling of patient’s tumors was assessed using an in-house next generation sequencing panel targeting genetic alteration in 571 genes relevant in cancer (DRAGON panel, [30]). The DRAGON panel retrieves both mutations and CNV. The DRAGON panel was combined to the DIAMOND assay [29] to perform multimodal ctDNA profiling of plasma samples (**Figure 1B-C**). DIAMOND profiles DNA methylation and the copy number of hominoid LINE-1 (L1PA) retrotransposons, which are typically altered in cancer, and have been shown to be highly promising multicancer biomarkers [29]. Combining both methods enabled the simultaneous detection of single nucleotides variants (SNVs) within the 571 gene tested, the profiling of L1PA DNA methylation and the evaluation of CNV over the 2.7 Mb of the gene panel and the 97.9 Mb covered by L1PA targets (**Figure 1C-D**).

### Landscape of molecular alterations detected from tumor and plasma DNA at baseline in SHIVA02 patients treated with targeted therapy

The most frequently altered genes in tumor samples were *TP53*, *PIK3CA*, *CDKN2A/B*, and *KRAS* (**Figure 2A**) with additional recurrent alterations observed also in *PTEN*, *ERBB2*, *APC*, *DNMT3A*, *BRAF*, *BRCA1*, *CCND1*, *ESR1*, *KMT2C*, *MAP2K4*, *NF1*, and *NRAS*. Variant types included missense, truncation, splice-site, promoter-region variants, in-frame deletions, homozygous deletions, and amplifications. At baseline, 62.5% of patients (20/32) had detectable ctDNA based on mutation profiling, with a median of 3 mutations per patient (**Figure 2A, Table S2**). The most frequently found SNVs in plasma DNA are those detected in the *TP53*, *PIK3CA*, and *KRAS* genes. Among patients with detectable ctDNA, 80.6% of alterations were shared between tumor and ctDNA (58/72), 12.5% were tumor-specific (9/72), and 6.9% were ctDNA-specific (5/72), highlighting heterogeneity in shedding and molecular detectability across patients (**Table S2, Figure 2A**).

**Figure 2.**
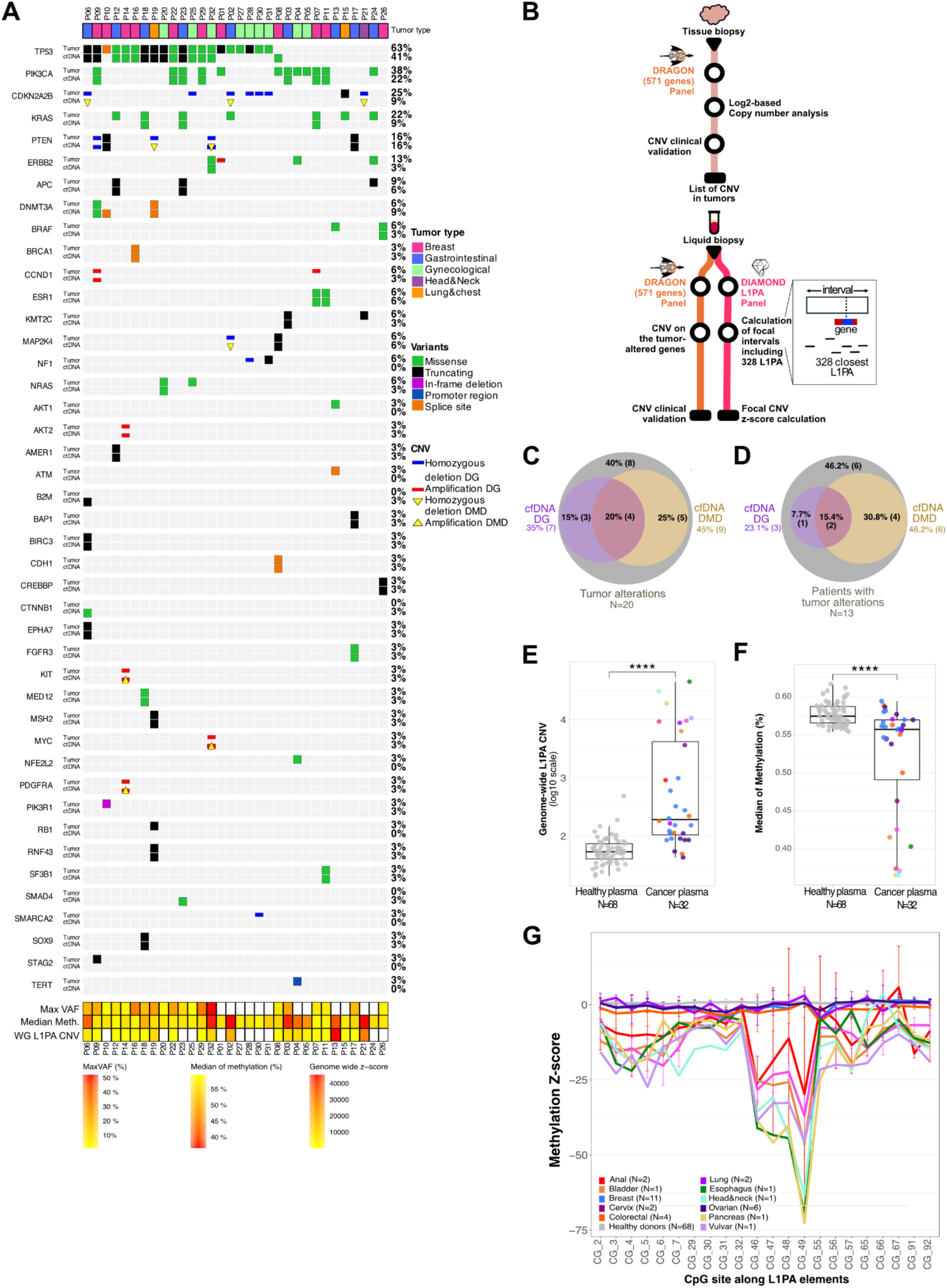
Landscape of molecular alterations detected from tumor and plasma DNA at baseline in SHIVA02 patients treated with targeted therapy. **A**. Oncoprint representation of the most frequent mutations detected in tumors vs plasma DNA. Alterations are organized per gene ranked by decreasing mutant frequencies with alterations found in tumors on top of corresponding alterations found in baseline plasma DNA. DRAGON gene-based profiling retrieves both mutations and CNV alterations while DIAMOND L1PA-based profiling retrieves L1PA hypomethylation and CNV. CNV detected with DIAMOND is highlighted with yellow triangles. Global parameters – maximum of variant allele frequencies (MaxVAF), median of L1PA DNA methylation (Median Meth.) and whole-genome L1PA CNV scores (WG L1PA CNV) are represented for each patient at the bottom of the oncoprint. **B.** Description of the DRAGON and DIAMOND methods used for focal CNV **C-D.** Venn diagram showing overlap between tumor CNV and CNV detected in cfDNA by alterations (**C**, 12/20 CNV alterations, 60 %) or patients (**D**, 8/13 patients, 61.5 %). DG: DRAGON, DMD: DIAMOND. **E-F.** Boxplot of L1PA genome-wide CNV scores (**E**) and median methylation levels (**F**) in 68 healthy donors (HD) and the 32 baseline cancer plasma samples, colored by cancer type. Statistical comparisons were performed using the Wilcoxon test. **G.** Differential methylation levels in healthy samples (N = 68) and patients for each type of cancer represented as z-scores along L1PA CpG sites. Error bars represent standard deviations.

To compare rates and specificities of CNV detected with DRAGON and DIAMOND, we implemented a pipeline to detect focal CNVs using DIAMOND, which previously had only been used at the genome wide level [29] (**Figure 2B**, Methods section and **S1A**). A total of 20 CNVs were identified in tumors of 13 patients (**Figure 2C-D, Table S2**). Among these, 12 (60%) were detected in matched ctDNA using at least one analytical method (**Figure 2C**), corresponding to 8/13 patients (**Figure 2D**). DRAGON and DIAMOND showed complementary performance: 7/12 (58%) plasma-detected CNVs were identified by DRAGON and 9/12 (75%) by DIAMOND, with most alterations (8/12) captured by only one method (**Figure 2A, 2C**). Deletions were more frequent than amplifications in tumor samples (65% vs. 35%, **Figure S1B**) and their detection rates in ctDNA differed accordingly. Respectively, 71 % (5/7) of the amplifications and 54 % (7/13) of the deletions were also detected in ctDNA (**Figure S1C**). Amplifications were more reliably detected by DRAGON (5/7, 71%), whereas DIAMOND showed superior sensitivity for deletions (6/7, 85% of plasma-detected deletions). Notably, concordant detection of CDKN2A/2B deletions in ctDNA was observed in half of the affected patients, exclusively through the DIAMOND approach. All PTEN alterations identified in tumor tissue were detected in plasma when combining both methods.

Next, to assess general aneuploidy, we computed genome-wide L1PA CNV scores, combining chromosome arm levels (**Table S3**) as previously performed with DIAMOND [29]. As expected, scores in baseline cancer plasmas are significantly higher than in healthy controls (**Figure 2E**). In parallel, we evaluated median L1PA methylation levels and observed significant hypomethylation in cancer patients (**Figure 2F**). This hypomethylation was detected across the entire length of L1PA elements (**Figure 2G**). Importantly, significant hypomethylation along the L1PA element is observed in cancer types that were not involved in the DIAMOND method’s optimization [29] (**Figure S1D**) underscoring its robust pan-cancer relevance. Indeed, we confirmed that tumors from these new cancer types analyzed (anal – P03 and P17, pancreas – P02, esophageal – P13, head and neck – P21, and bladder – P06) displayed clear hypomethylation in an independent public dataset [31] (**Figure S1E**).

### Combining mutational and L1PA profiling increases ctDNA detection rate at baseline

Next, we have evaluated the detectability of ctDNA at baseline based on the three different molecular parameters: maximum variant allele frequencies (MaxVAF), L1PA hypomethylation rates (MethP_Cancer_) and L1PA CNV scores. To do so, we have determined detection thresholds for each analytical approach (see Methods). The detection threshold for SNVs was set at MaxVAF ≥ 1%, corresponding to the clinically validated limit of detection of the DRAGON assay. The threshold for L1PA DNA methylation was defined to maximize the sensitivity at 98% specificity in a previously studied independent cohort [29] and the threshold for genome-wide L1PA CNVs was determined in our previous study [29] (**Figure 3A-B**). When applying these thresholds using each parameter individually, ctDNA detectability reached 62.5 % (20/32) with mutation profiling, 65.6 % (21/32) with L1PA CNV and 90.6 % (29/32) with L1PA methylation analysis alone (**Figure 3C**, **Table S4**). We further analyzed the relationship between mutations and L1PA alterations. We compared the mutation MaxVAF with L1PA methylation levels, or with L1PA genome-wide CNV scores for each patient at baseline and did not observe any correlation (**Figure 3D**, **S2A**). When removing samples with MaxVAF below 1 %, we revealed a slight correlation for the methylation metrics and good correlation for the CNV (**Figure S2B**). This suggests that the genomic and epigenetic features provide distinct and potentially complementary information reflecting different aspects of tumor genomes, in particular for samples with low mutant allele frequencies.

**Figure 3.**
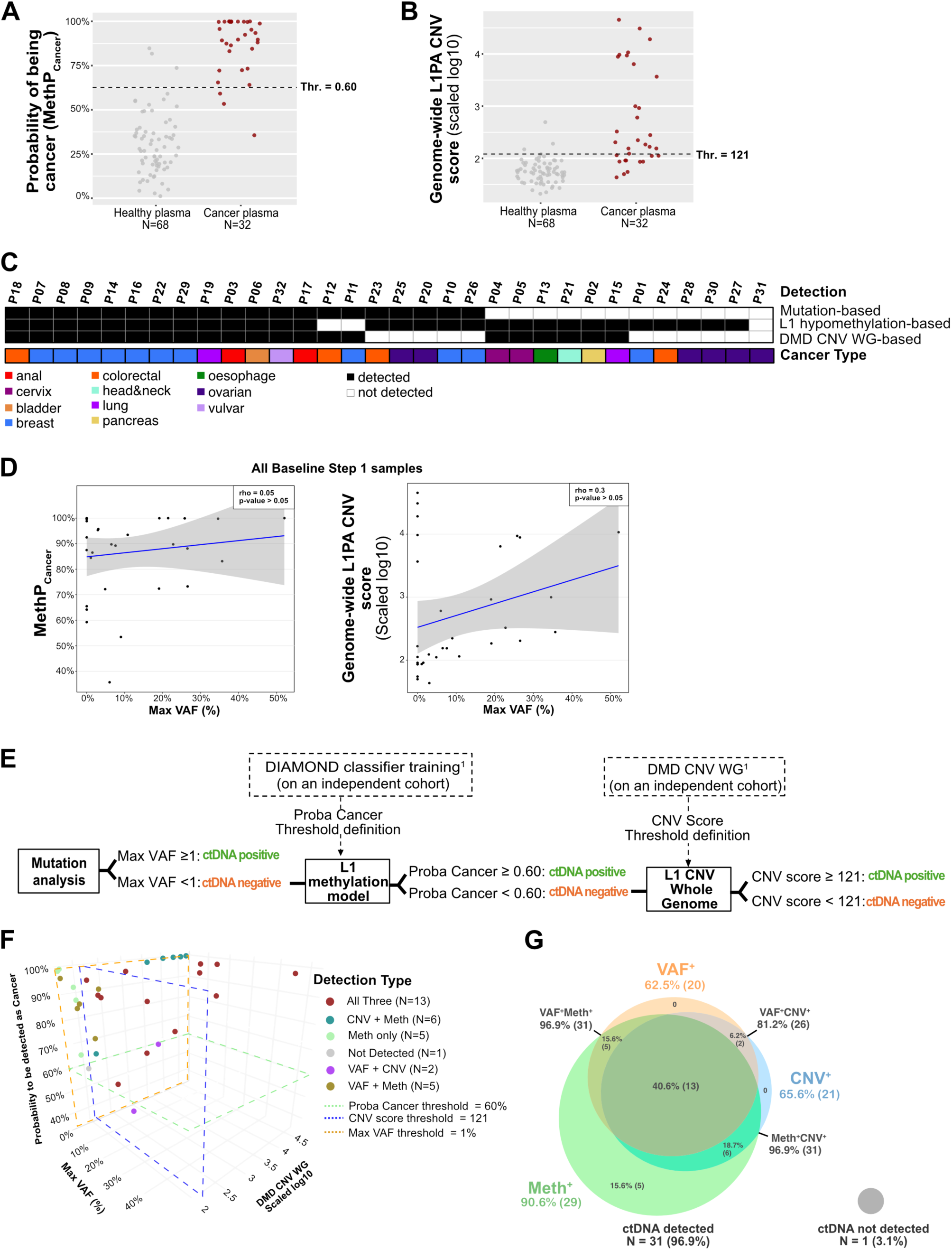
Combining mutational and L1PA profiling increases ctDNA detection rate at baseline. A-B. Methylation-based model probability of being cancer (MethP_Cancer_, **A**) or genome-wide L1PA CNV score (**B**) in 68 healthy donors and 32 baseline cancer plasma samples. The threshold for L1PA DNA methylation was defined to maximize the sensitivity for a 98% specificity on an independent cohort previously studied [29] and genome-wide L1PA CNV was determined in our previous study [29]. Both are indicated by horizontal grey dotted lines. Thr.: threshold. **C**. Heatmap showing the detection results for each analysis parameter for each patient, according to cancer type. Overall, 31/32 patient were detected (96.9%). **D.** Correlation analysis for highest variant allele frequency (MaxVAF) with the methylation model cancer probability (MethP_Cancer_, left panel) or with L1PA CNV scores (right panel) of the all-baseline samples. **E.** Three-step detection model workflow. CtDNA positivity based on mutation profiling was defined as a MaxVAF ≥ 1%. L1PA hypomethylation positivity was defined using the cancer probability threshold of 0.60. Genome-wide L1PA CNV positivity was defined using a threshold of 121. Both L1PA thresholds were previously established in an independent healthy-versus-tumor cohort [29]. **F.** 3D dot plot highlighting patients detected with the 3 types of alterations (MaxVAF_threshold_ ≥ 1%, MethP_Cancer threshold_ ≥ 60%, L1PA CNV score_threshold_ ≥ 121). **G.** Venn diagram of the number of patients detected by each method at baseline.

To evaluate the complementarity of these alterations for ctDNA detection, we applied a three-step detection model integrating the three molecular parameters. Mutation status was evaluated first, followed by L1PA hypomethylation and L1PA CNV for patients not detected at the previous steps (**Figure 3E**). The joint distribution of baseline MaxVAF values, MethP_Cancer_, and genome-wide L1PA CNV scores showed that several patients were negative by mutation-based detection but detected by L1PA hypomethylation and/or CNV analysis (N = 11, **Figure 3C** and **3F-G**). Indeed, the combined detection model increased ctDNA detectability to 96.9% (31/32, **Figure 3G**). Notably, across cancer types, 13 patients were identified by all three parameters, 13 by two parameters, and 5 by L1PA hypomethylation alone (**Figure 3C**). Patient P31 was not detected with the combined model. We compared this patient with patients showing similar molecular profiles but detected with the model. Patients P01, P24, P28 and P30 displayed MaxVAFs < 1%, L1PA CNV < 121, and a median methylation level as low as that of P31 (**Figure 2A bottom**) but were nevertheless detected by our model, as all exceeded the probability threshold driven by L1PA hypomethylation (MethP_Cancer_ : P01 = 92.4%, P24 = 99.7%, P28 = 87.5%, P30 = 99.7%; **Table S2**). Although median methylation levels were comparable, individual CpG hypomethylation was more pronounced, especially in P01, P24 and P30, compared to P31 (**Figure S2C**), which likely explains why P31 was not detected. These findings further support the notion that the methods interrogate partially distinct tumor-derived signals, thereby providing complementary rather than redundant information. Collectively, these results support the added value of multimodal ctDNA profiling, suggesting that the integration of genetic and tumor-naive (epigenetic alterations enhances sensitivity and provides a more comprehensive assessment of tumor-derived signals at baseline.

### CtDNA detected with both mutations and L1PA alterations reflect response to treatment and can detect relapse early

Next, we studied the potential of this combined model for disease monitoring. Of the 32 patients included, 21 had at least one follow-up sample (**Figure 1B**). To better understand their interplay, we assessed correlations between the three parameters across longitudinal samples. We observed a low positive correlation between mutations and L1PA methylation levels (rho = 0.36, p < 0.05) and with genome-wide L1PA CNV score (rho = 0.24, p < 0.05, **Figure 4A**). Correlations are both reinforced when focusing on samples with a MaxVAF above the 1% detection threshold (**Figure S2D**). This suggests that, while these three molecular parameters capture partially distinct tumor-derived signals, their dynamic evolution during treatment is broadly concordant, reflecting shared underlying changes in tumor burden, even in the absence of a linear correlation at specific timepoints. Longitudinal monitoring of ctDNA during treatment revealed patient-specific dynamics and temporal variations were observed across all three molecular parameters. Overall, periods of disease progression coincided with higher MaxVAF, MethP_Cancer_, and L1PA CNV scores, while lower or undetectable values were observed during disease control (**Figure 4B**).

**Figure 4.**
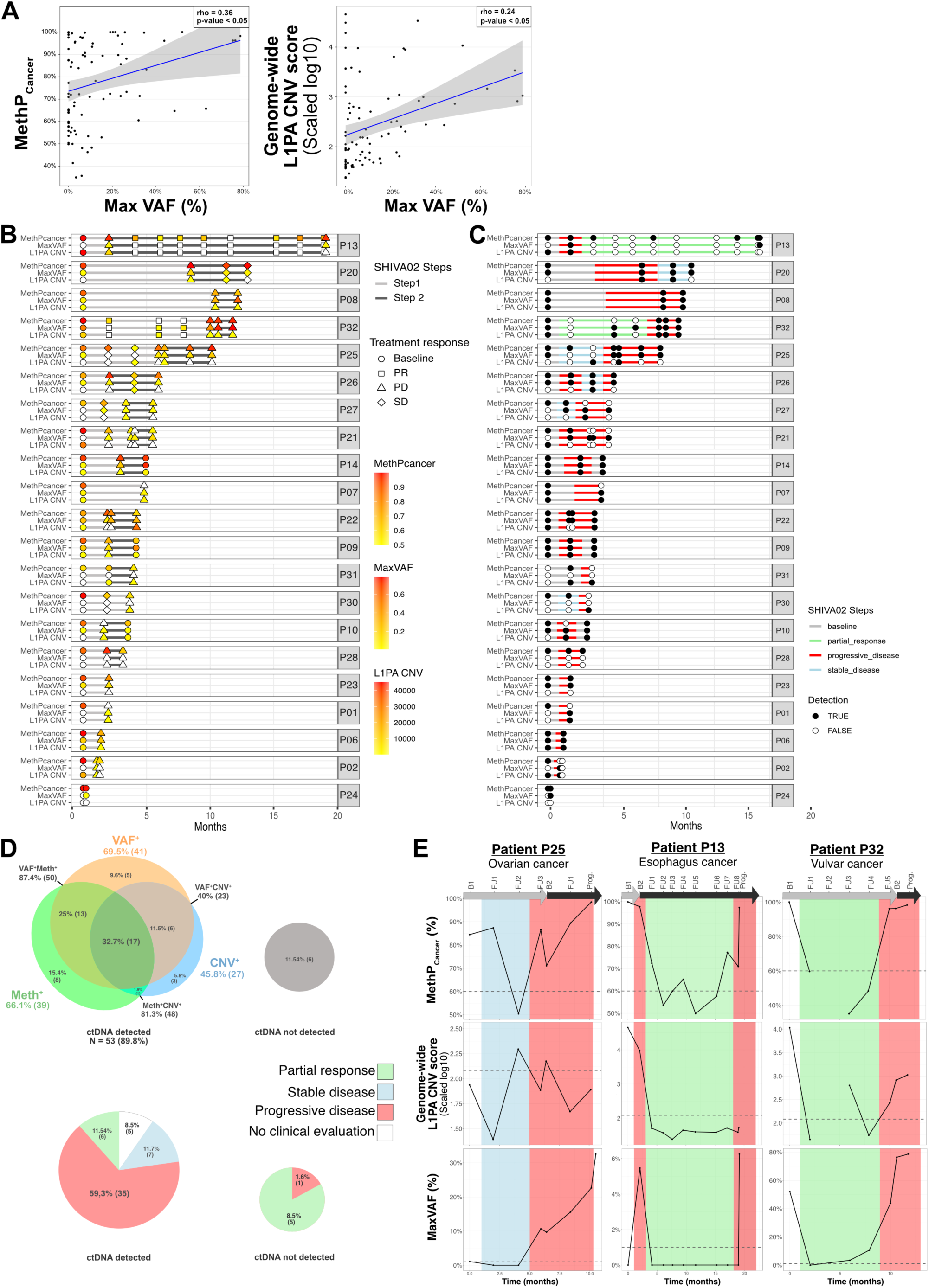
CtDNA detected with mutations and L1PA alterations reflect response to treatment and can detect relapse early. **A.** Correlation analysis of MaxVAF with MethP_Cancer_ (left panel) or L1PA CNV (right panel) in samples from follow-up timepoints (N=91). **B-C.** Swimmer plots of patients with more than one timepoint over time (months), displaying the quantitative values of the three metrics (MethP_Cancer_, MaxVAF, and L1PA CNV) (**B**) and their corresponding ctDNA detection status in a binary format (**C**, black: detected, white: not detected). **D.** Venn diagram of the number of follow-up samples with ctDNA detected or not detected, classified by each metric (top panel) or by treatment response status (bottom panel). **E.** Dynamics of the three metrics through disease evolution (green: partial response, blue: stable disease, red: progression) for three selected patients (see **Figure S3** for all the patients). B: Baseline, FU: follow-up time point. The 3^rd^ timepoint (FU2) of P32 misses DIAMOND metrics as its library failed quality control.

We next evaluated ctDNA detection across all samples over time using the three metrics and analyzed the presence or absence of a signal in relation to the response to treatment (**Figure 4C**). While L1PA methylation levels proved to be an important parameter in the 3-step detection model at baseline, the MaxVAF is a key factor for disease monitoring, enabling the detection of 69.5% (41/59) of follow-up samples on its own (**Figure 4D**). Overall, we observed higher MaxVAFs in follow-up samples than at baseline (**Figure S2E**). However, applying the 3-step model increased the detection rate to 89.8% (53/59) of the follow-up samples regardless of the treatment response status. More precisely, 36/37 (97.3 %) of follow-up samples were detected at progression, 7/7 (100 %) at stable disease, 3/11 (27.3 %) at partial response and 4/4 (100 %) with no clinical information. Among the 11/59 (18.6%) undetected follow-up samples, 8/11 (72.7 %) were collected during partial response, consistent with reduced tumor burden and shedding under effective treatment (**Figure 4D**). Only one patient, P02, was not detected at progression. This patient displayed MaxVAF values equal to or below the detection threshold throughout disease monitoring, suggesting persistently low tumor burden and shedding even during clinical progression (**Figure S3**). Patient P31, who was just below L1PA thresholds at baseline, displayed L1PA reactivation signatures rising above detection thresholds during monitoring, enabling ctDNA detection over time. (**Figure 4B-C**, **S3**).

We next analyzed more precisely the ctDNA dynamics in relation to the response to treatment in three patients who had 7-10 timepoints sampled over time (**Figure 4E**). Overall, longitudinal dynamics in MethP_Cancer_, L1PA CNV score, and MaxVAF displayed similar trends for each patient. More importantly, we observed that changes in one or more cfDNA parameters preceded clinical evidence of progression (**Figure 4E**, **S3**). Notably, in patient P32, with vulvar cancer, the mutation-based approach detected a rise in MaxVAF (FU3, FU4) approximately 4 months prior to clinical evidence of progression. P25, with ovarian cancer, displayed a rising L1PA CNV preceding mutations and methylation. The subsequent follow-up was negative but remained close to the detection threshold, and the patient then developed progressive disease. In P13, treated for esophagus cancer, a cancer reported with high LINE-1 reactivation [34], FU5 sample tested positive for MethP_Cancer_ parameter 2 months before the progression was observed clinically. This highlights the potential value of integrating multi-molecular analysis to estimate the earliest possible risk of relapse.

## Discussion

In this retrospective analysis of patients enrolled in the SHIVA02 precision-oncology trial, we assessed the value of a multimodal ctDNA profiling strategy combining genomic and tumor- naive epigenomic analyses for disease detection and monitoring in patients receiving molecularly matched targeted therapies. The three-step integrative model achieved a baseline ctDNA detection rate of 96.9%, substantially higher than any single modality alone, and longitudinal dynamics across all three parameters broadly mirrored treatment response and disease progression. Mutation-based ctDNA detection at baseline (65.6%) was consistent with previous reported rates in advanced solid tumors [11], [12]. Concordance between tumor tissue and plasma-derived mutations was partial, with most alterations shared between compartments but a subset detected exclusively in plasma or tumor tissue. Such discordance has been widely reported and likely reflects a combination of biological factors—including intratumoral heterogeneity, variable tumor DNA shedding, and clonal evolution under treatment pressure—as well as technical limitations inherent to sequencing-based approaches, including their analytical sensitivity for low-frequency variants, which may limit ctDNA detection even when tumor-derived DNA is present [12], [13]. While NGS offers the advantage of broad mutational profiling, more sensitive, targeted approaches have been shown to improve ctDNA detection in low-shedding settings, highlighting the complementary nature of different analytical strategies for disease monitoring [13].

CNV analysis further highlighted these challenges: 60% of tumor-derived CNVs were detectable in matched ctDNA using at least one or both analytical methods, confirming that CNV detection on its own remains challenging [12]. Stratification by CNV type revealed distinct detection patterns between amplifications and deletions, with method-dependent differences observed between the DRAGON and DIAMOND assays. These findings reinforce the view that no single analytical platform captures the full spectrum of tumor-derived genomic alterations in plasma and support the use of complementary analytical strategies.

A key contribution of this study is the integration of L1PA hypomethylation as an epigenetic biomarker of ctDNA. L1PA hypomethylation identified ctDNA in 90.6% of patients at baseline including cases with undetectable mutations and showed limited correlation with mutation- and CNV-based metrics — supporting the hypothesis that these approaches capture distinct biological dimensions of tumor activity. This finding is consistent with accumulating evidence indicating that global DNA hypomethylation represents a widespread and stable hallmark of cancer that can be sensitively detected in circulating cell-free DNA [17]. Extending the approach originally developed in Michel *et al.* [29], the present study further demonstrates, the applicability of retrotransposon hypomethylation-based ctDNA detection across several new tumor types including bladder, anal, esophageal, cervical, pancreatic, and head and neck cancers, supporting its broader utility as a sensitive and specific biomarker for the noninvasive detection and monitoring of a wide spectrum of malignancies.

Importantly, the limited correlation observed between mutation-based, CNV-based, and methylation-based ctDNA metrics suggests that these approaches capture distinct biological dimensions of tumor activity. Whereas mutation-based metrics primarily reflect clonal tumor populations, L1PA hypomethylation likely captures a more global tumor-derived signal. Their combined use substantially increased overall ctDNA detectability, highlighting the added value of integrated genomic and epigenomic profiling.

Longitudinal analyses demonstrated that dynamic changes in variant allele frequencies, L1PA methylation levels, and CNV patterns closely mirrored treatment response and disease progression. In some patients, changes in one or more ctDNA metrics were observed before radiological or clinical progression, supporting the potential utility of multimodal ctDNA monitoring for early detection of therapeutic resistance. These findings are consistent with previous studies showing that ctDNA dynamics can anticipate disease progression across multiple tumor types and treatment contexts, including lung, breast, and colorectal cancers [13], [14], [15], [16]. Notably, our observations extend these findings to a pan-cancer population treated with heterogeneous targeted therapies, supporting the broader applicability of this monitoring strategy in precision oncology. This multimodal profiling of ctDNA could prove particularly useful for a wide range of cancer types, including tumors with low mutational burden — such as sarcomas or pediatric tumors — for which methylation signals would be helpful in detecting ctDNA.

Several limitations of this study should be discussed. The retrospective design and limited sample size restrict statistical power and limit the interpretation of the results across tumor types. All patients had advanced and / or metastatic disease, a clinical context known to be associated with higher levels of ctDNA shedding, which may have facilitated ctDNA detection and influenced the observed detection rates, although low-shedding metastatic tumors have also been documented [30]. In addition, tumor-type heterogeneity and diversity of targeted therapies preclude tumor-specific conclusions. Finally, discordant SNV calls between tissue and plasma highlight ongoing technical and biological challenges in ctDNA analysis that warrant further investigation in larger, prospective cohorts. An additional practical consideration concerns the economic feasibility of multimodal ctDNA profiling. Comprehensive Genomic Profiling panels such as DRAGON already represent a significant cost per analysis, and the addition of methylation profiling further increases the overall analytical burden. The cost-effectiveness of such combined approaches will therefore need to be formally evaluated prior to consideration for routine clinical implementation. Ultimately DRAGON and DIAMOND targets could be combined in a single panel to optimize the analysis and reduce the costs. Taken together, these findings support the prospective evaluation of multimodal ctDNA profiling in larger, homogeneous cohorts to determine whether the integration of genomic and epigenomic biomarkers can meaningfully inform therapeutic decision-making in precision oncology.

## Conclusion

This study demonstrates that integrating mutation-based profiling, CNV analysis, and tumor- naive LINE-1 hypomethylation detection substantially improves ctDNA detection and provides complementary insights into tumor dynamics in patients with advanced cancer receiving matched targeted therapies. These findings support the use of multimodal ctDNA approaches for molecular monitoring in precision oncology and provide a rationale for prospective validation in larger patient cohorts.

## Data Availability

Analyses are based on processed next generation sequencing data which are provided in supplemental tables.
The sequencing data will be available upon reasonable request to the authors and accessible via EGA.

## Acknowledgments

We thank the patients and their families for their participation and trust in the SHIVA02 trial. We thank the medical, nursing, and clinical research staff of Institut Curie involved in the conduct of the SHIVA02 trial, including patient care, sample collection, and data management. This work was supported by grants from the European Research Council (ERC-StG EpiDetect) and Rennes Metropole (Scientific Installation Grant) of which C. Proudhon was recipient, and a grant from MSD Avenir Foundation of which C. Le Tourneau was recipient.

## Declaration of interests

C. Proudhon is co-author of a patent related to the DIAMOND method (PCT/EP2023/074092) “Sensitive and Specific Determination of DNA Methylation Profiles”. M. Kamal reports personal fees from Roche outside the submitted work. C. Dupain reports personal fees from Astrazeneca outside the submitted work. C. Le Tourneau reports personal fees from MSD, Bristol Myers Squibb, Merck, AstraZeneca, Celgene, Seattle Genetics, Roche, Novartis, Rakuten, Nanobiotix, and GSK outside the submitted work.

## Materials and Methods

### Samples collection

Cancer patients’ samples and data were obtained from 32 patients included in SHIVA02 clinical trial (NCT03084757), promoted by Institut Curie (Paris, France), with druggable molecular alterations detected in their tumor and treated by matched therapy in the frame of the trial (Du Rusquec *et al.* under review). All patients had recurrent or metastatic solid tumors and underwent molecular profiling as part of the SHIVA02 study. Written informed consent was obtained from all participants, and the SHIVA02 trial was conducted in accordance with institutional guidelines and the Declaration of Helsinki. Tumors were collected at study inclusion by core-needle biopsy. Tumor cellularity was assessed by a pathologist, and only biopsies containing more than 30% tumor cells were frozen and then used for DNA extraction. Plasma samples were collected at baseline and longitudinally during the study, approximately every two months and at the time of disease progression.

Healthy plasma were collected from blood of healthy donors through the French blood establishment under French and European ethical practices to be used as control in addition of 60 healthy plasma from Michel et al., for the DIAMOND method [29]. Healthy peripheral blood samples were collected in Cell-Free DNA BCT® tubes and processed by sequential centrifugation to isolate plasma, which was aliquoted and stored at −80 °C until cell-free DNA extraction.

### DNA extraction

Tumor DNA was extracted from frozen biopsy specimens using a phenol–chloroform–isoamyl alcohol extraction method (Invitrogen), according to the manufacturer’s instructions. Extracted DNA was quantified using both spectrophotometric measurement (NanoDrop™ ND-2000, Thermo Scientific) and fluorometric quantification (Qubit™, Life Technologies, Carlsbad, CA, USA). DNA integrity was assessed by agarose gel electrophoresis. DNA samples were subsequently stored at −20 °C until library preparation and downstream molecular analyses. Cell-free DNA was extracted from plasma samples using the QIAsymphony Circulating DNA Kit (Qiagen) or the Maxwell RSC ccfDNA LV Plasma kit, following the manufacturer’s recommendations. The impact of the extraction method on downstream results has been shown to be negligible, as previously reported [29]. Extracted cfDNA was subsequently used for circulating tumor DNA analyses.

### Targeted NGS analysis using the DRAGON panel

A comprehensive genomic profiling (CGP) assay was performed using the DRAGON panel (Detection of Relevant Alterations in Genes involved in Oncogenetics by NGS), commercially available as the SureSelect CD Curie CGP assay (Agilent). This targeted panel covers key oncogenic drivers and tumor suppressor genes and enables the detection of single-nucleotide variants, small insertions / deletions, and copy-number alterations. Libraries were prepared and sequenced according to the manufacturer’s recommendations, and data were processed using the standardized DRAGON bioinformatics pipeline for variant calling and annotation as previously described [11], [31]. Alterations were classified according to current clinical guidelines [10]. For plasma analyses, samples were considered ctDNA-positive when at least one somatic alteration was detected with a maximum variant allele frequency (MaxVAF) of 1% or higher. The 1% VAF threshold corresponds to the clinically validated limit of detection of the DRAGON assay, determined through a series of analytical validation experiments. This cutoff ensures reliable SNV detection while minimizing false positives due to sequencing noise.

### Targeted NGS analysis using the DIAMOND assay

#### L1PA DNA methylation analysis

CfDNA samples were profiled using the DIAMOND assay (Detection of Long Interspersed Nuclear Element Altered Methylation ON plasma DNA) [29]. Briefly, targeted bisulfite sequencing libraries were prepared as previously described, except that amplicon 2 was excluded due to its high variability. Sequencing was performed on an Illumina NextSeq 2000 or NovaSeq platform (PE 30 bp, 170 bp). CpG methylation calling and haplotype extraction were conducted as described previously [29]. The resulting methylation profiles were evaluated using the “All cancer model”, a Random Forest classifier trained on the average methylation levels across 22 CpG sites in a discovery cohort comprising 6 cancer samples, as described in Michel *et al.* [29]. Changes in L1PA methylation compared with healthy individuals was calculated with the following z-score formula:

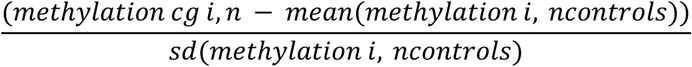

with *i* = a given L1PA CpG site, *n* = a given sample, and n *controls* = a set of 63 healthy plasmas from the discovery cohort of Michel *et al.* [29].

#### Public dataset to compare L1PA hypomethylation in new cancer types

To evaluate L1PA hypomethylation in cancer types not included in the DIAMOND method’s optimization previously published, we analyzed data from a recent study profiling cell-free DNA (cfDNA) methylation using long-read Oxford Nanopore Technologies (ONT) sequencing across tumor and adjacent healthy tissues [32](**Figure S1D**). Methylation frequencies from 15 tumor samples across five cancer types and 40 healthy samples reported by O’Neill *et al.* were retrieved. Using RepeatMasker with the hg38 reference genome, we retrieved L1PA CpG sites targeted by the DIAMOND assay and aligned them on the *L1HS* Repbase sequence. This was then intersected with the ONT-derived CpG methylation frequencies data using BEDTools. Methylation values were subsequently averaged by CpG position along the *L1HS* consensus sequence. All processing and analysis steps described above were implemented in ATHENA (v0.01) [33]. L1PA hypomethylation was calculated using the z-score with 15 tumor-adjacent healthy tissue samples serving as the reference control group.

#### Copy number variations analysis based on L1PA sequence profiling

Genome-wide CNV analysis was performed as described previously [29] using the same computational pipeline and reference cohort. Genome-wide z-scores were computed as the sum of squared chromosome-arm z-scores. For focal CNV analysis, interval optimization was done by increasing progressively the number of ‘probes’ analyzed for CNV detection. We screened intervals covering 15 to 400 L1PA targets around the altered gene of interest (using RepeatMasker hg38 annotations). This corresponds to intervals from 172.7Kb to 23.2 Mb. For each interval, normalized read counts were calculated per sample and z-scores were computed using the same formula applied at the chromosome-arm level for genome-wide scores. Significance of focal CNV z-scores was assessed using a two-sided threshold derived from a Student’s t-distribution, with degrees of freedom defined by the number of healthy donor reference samples used for z-score calculation - 1. Intervals maximizing the number of detected alterations in cfDNA, compared to the tumor, included 328 L1PA targets (**Figure S1C**). This corresponded to intervals from 4.9 Mb to 18 Mb depending on the region analyzed.

#### 3-steps classification for sample labelling

Sample classification was based on a three-step model. Samples with a maximum VAF (MaxVAF) ≥ 1% in the DRAGON assay were classified as cancer. Remaining samples were evaluated using the methylation-based model probability of being cancer (MethP_Cancer_) that maximized the sensitivity for a 98% specificity on an independent cohort described in our previous study [29]. Samples with a probability exceeding the predefined threshold were classified as cancer. For samples below this threshold, genome-wide z-scores were used as a final decision criterion using the genome-wide z-score threshold of 121 already determined in our previous study [29]. Classification rules were defined as follows: samples with max VAF ≥ 1% were classified as cancer; otherwise, samples were classified as healthy if their ProbaCancer ≤ 0.60 and genome-wide z-score ≤ 121, and as cancer if ProbaCancer > 0.60 or genome-wide z-score > 121 (**Figure 3C**).

### Statistical analyses

All correlations were assessed using Spearman’s rank correlation, and comparisons between healthy and cancer plasma were performed using the Wilcoxon rank-sum test.

## Supplementary materials

This includes:

**Figure S1.**
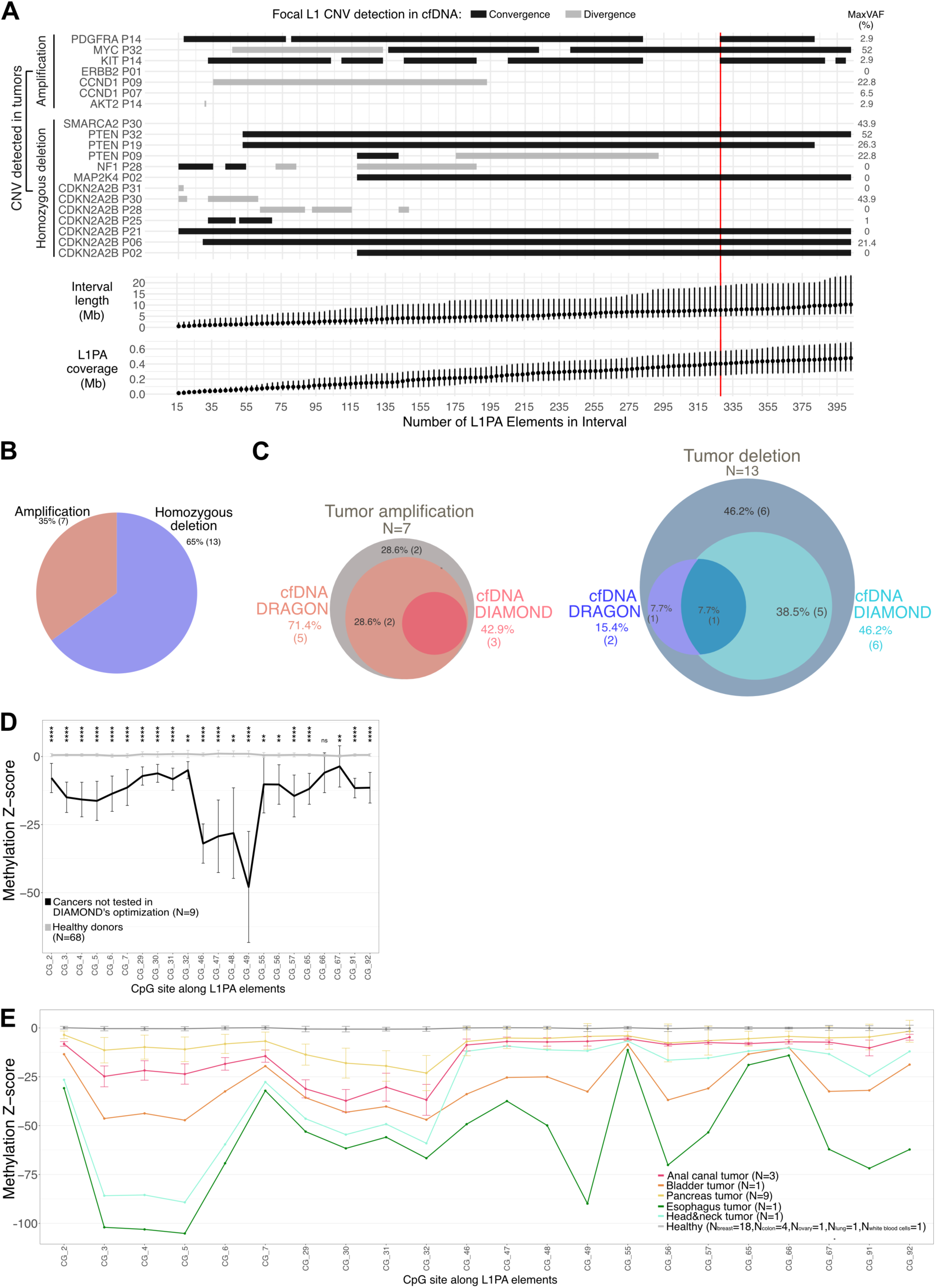
**A.** Optimization of focal copy number variant (CNV) detection across different intervals with a varying number of L1PA targets. Intervals ranging from 15 to 400 L1PA targets were screened for focal CNV detection in cfDNA. CNVs were detected based on the number of L1PA targets within the range, per gene and patient with tumor alterations. For each L1PA interval, the range of median sizes (4.9–18 Mb) and the L1PA coverage (247.8–571.7 kb) are shown. **B.** Pie chart showing the distribution of insertions and deletions detected using at least one of the two profiling techniques. **C.** Venn diagram of the number of tumor amplifications (left panel) or tumor deletions (right panel) detected by each method in cfDNA. **D.** Average differential methylation levels in healthy samples (N = 68) and patients not studied by Michel *et al.*, represented as z-scores along L1PA CpG sites (N = 9 – anal, pancreas, esophageal, head and neck, bladder, cervix and vulvar cancers). Statistical significances were computed using Mann–Whitney U test on methylation z-score. Error bars represent standard deviation. **E.** Differential methylation levels in healthy tissues and tumor samples matching cancer types analyzed in **D**. from O’Neill *et al.* [31]. Error bars represent standard deviation.

**Figure S2.**
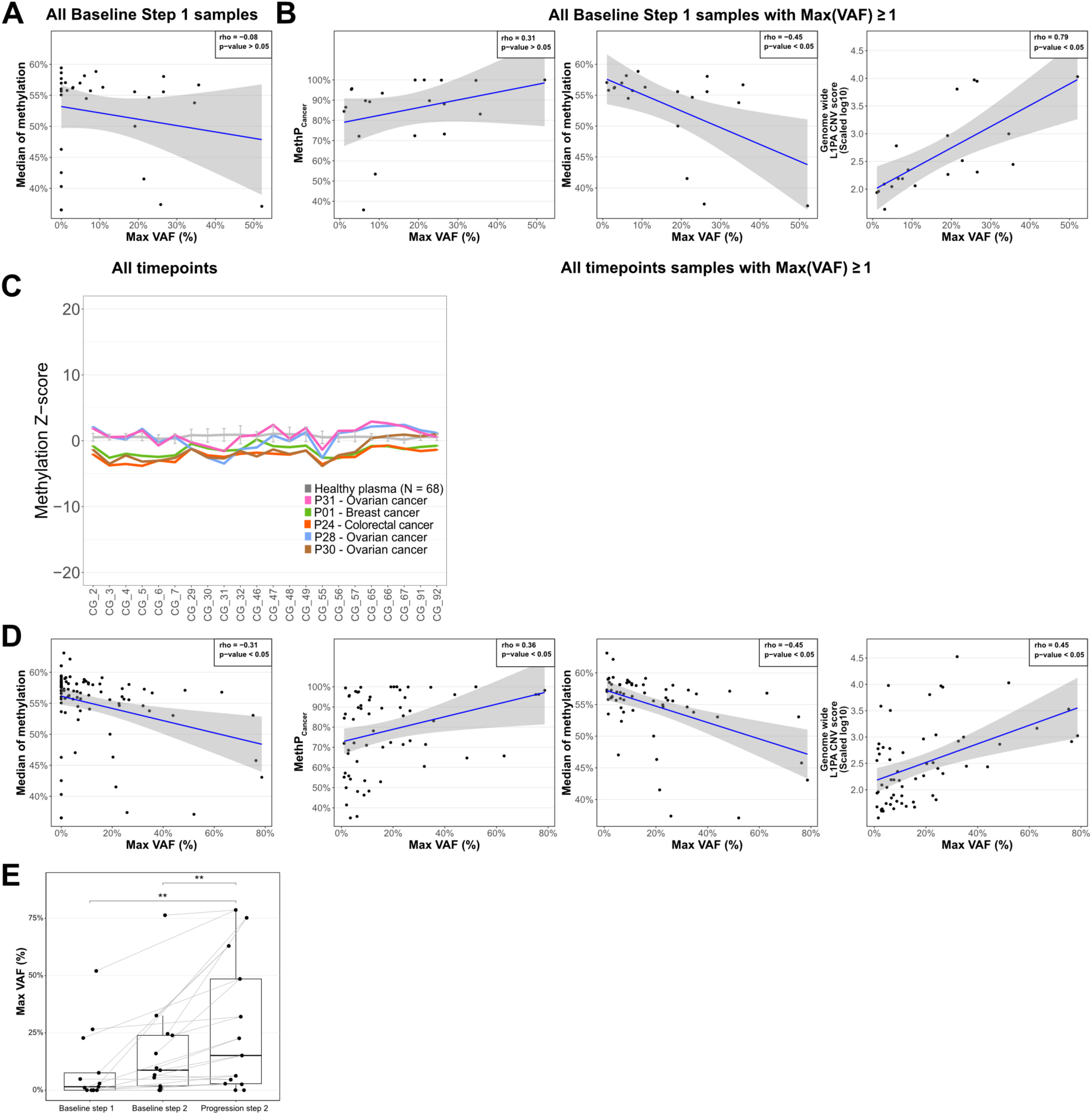
**A-B.** Correlation analysis for highest variant allele frequency (MaxVAF) with the median of L1PA methylation for all samples (**A**) or with MethP_Cancer_ (**B,** left panel), or with median of L1PA methylation (**B,** middle panel) or with L1PA CNV scores (**B**, right panel) of baseline samples, excluding samples with MaxVAF < 1 %. **C.** Comparison of differential methylation between samples detected (P24, P01, P28 and P30) and not detected (P31) by L1PA hypomethylation and healthy plasma (N=68). Error bars represented standard deviation. **D.** Correlation analysis for highest variant allele frequency (MaxVAF) with the median of L1PA methylation for all follow-up and baseline samples (left panel) or with MethP_Cancer_ (2^nd^ panel), median of L1PA methylation (3r^d^ panel), or with L1PA CNV scores (4^th^ panel) for all follow-up and baseline samples, excluding timepoints with MaxVAF < 1 %. **E**. Paired comparison boxplot of the highest variant allele frequency (MaxVAF) between Baseline Step 1, Baseline Step 2 and Progression Step 2 samples. Statistical significances were computed using the signed paired Mann–Whitney U test.

**Figure S3.**
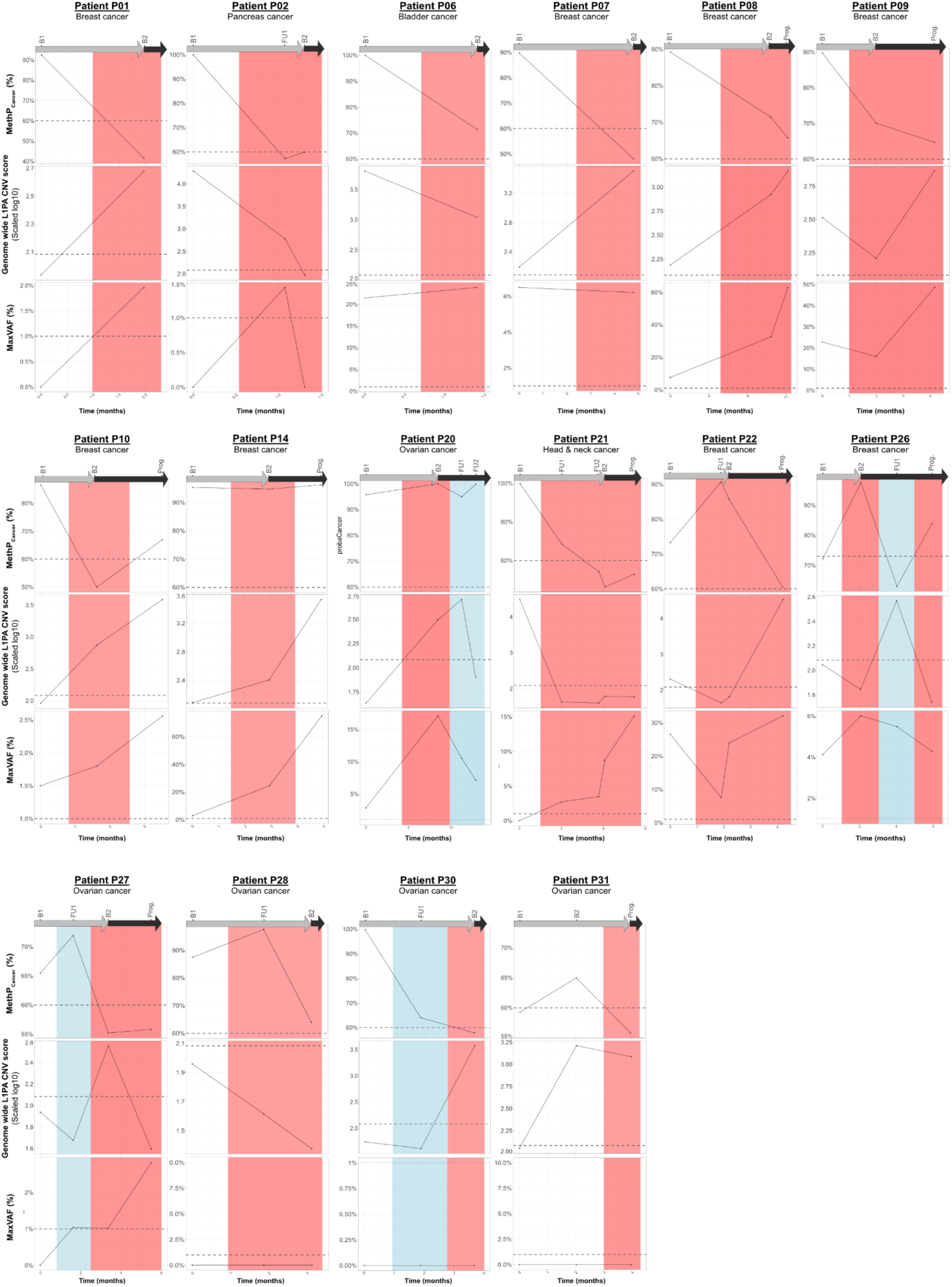
Longitudinal evolution of the three metrics: mutation-based ctDNA levels (MaxVAF), L1PA-methylation-based cancer probabilities (MethP_Canc_er), genome-wide L1PA CNV scores (Genome-wide L1PA CNV score) in patients with two or more follow-up samples, in relation to their clinical disease evaluation (White: Baseline steps, Blue: Stable disease. Red: Progressive disease).

Tables S1 to S5 as separated Excel file

## Notes

### Clinical Trial

NCT03084757

### Author Declarations

Written informed consent was obtained from all participants, and the SHIVA02 trial (NCT03084757) was conducted in accordance with institutional guidelines and the Declaration of Helsinki. This study was approved by the Curie Institutional Board under approvals INT210320 and DATA210182.

### Summary of Updates

Result section as been amended and updated to clarify; author list has been updated to better reflect the work done.

